# A digital self-care intervention for Ugandan patients with heart failure and their clinicians: User-centred design and usability study

**DOI:** 10.1101/2022.01.13.22268796

**Authors:** Jason Hearn, Sahr Wali, Patience Birungi, Joseph A. Cafazzo, Isaac Ssinabulya, Ann R. Akiteng, Heather J. Ross, Emily Seto, Jeremy I. Schwartz

**Affiliations:** Institute of Biomedical Engineering, University of Toronto, Toronto, ON, Canada; Centre for Global eHealth Innovation, Techna Institute, University Health Network, Toronto, ON, Canada; Institute of Health Policy, Management and Evaluation, Dalla Lana School of Public Health, University of Toronto, Toronto, ON, Canada; Department of Epidemiology and Biostatistics, Makerere University School of Public Health, Kampala, Uganda; Uganda Heart Institute, Mulago Hospital, Kampala, Uganda; Department of Medicine, Makerere University College of Health Sciences, Kampala, Uganda; Uganda Initiative for Integrated Management of Non-Communicable Diseases, Kampala, Uganda; Ted Rogers Centre for Heart Research, Peter Munk Cardiac Centre, University Health Network, Toronto, ON, Canada; Department of Medicine, University of Toronto, Toronto, ON, Canada; Section of General Internal Medicine, Yale University School of Medicine, New Haven, CT, United States of America

## Abstract

**Background:** The prevalence of heart failure (HF) is increasing in Uganda. Ugandan patients with HF report receiving limited information about their illness, disease management, or empowerment to engage in self-care behaviors. Interventions targeted at improving HF self-care have been shown to improve patient quality of life and to reduce hospitalizations in high-income countries. However, such interventions remain underutilized in resource-limited settings like Uganda.

**Objective:** To develop a digital health intervention that enables improved self-care amongst HF patients in Uganda.

**Methods:** We implemented a user-centred design process to develop a self-care intervention entitled *Medly Uganda*. The ideation phase comprised a systematic scoping review and preliminary data collection amongst HF patients and clinicians in Uganda. An iterative design process was then used to advance an initial prototype into a fully-functional digital health intervention. The evaluation phase involved usability testing of the developed intervention amongst Ugandan patients with HF and their clinicians.

**Results:** *Medly Uganda* is a digital health intervention that is fully integrated within a government-operated mobile health platform. The system allows patients to report daily HF symptoms, receive tailored treatment advice, and connect with a clinician when showing signs of decompensation. *Medly Uganda* harnesses Unstructured Supplementary Service Data technology that is already widely used in Uganda for mobile phone-based financial transactions. Usability testing showed the system to be accepted by patients, caregivers, and clinicians.

**Conclusions:** *Medly Uganda* is a fully-functional and well-accepted digital health intervention that enables Ugandan HF patients to better care for themselves. Moving forward, we expect the system to help decongest cardiac clinics and improve self-care efficacy amongst HF patients in Uganda.

## Introduction

The prevalence of cardiovascular disease (CVD) is on the rise in Uganda^1^, with CVD now accounting for roughly 10% of deaths in the country^2^. This increasing prevalence of CVD places an immense burden on cardiac clinics, specifically given that African patients with CVD often present with various structural and hemodynamic complications, such as heart failure (HF), that require both acute and long-term management^3^. In the chronic phase of HF, patients require complex treatment plans involving a myriad of medications, lifestyle recommendations and self-care behaviours^4^. These management plans can be burdensome for both HF patients and clinicians, given the necessity for follow-up appointments to monitor patient health, the financial costs associated with medications, the need for adherence to stringent self-care instructions, and the overall impact of HF on patient quality of life^5^. Ugandan patients with HF describe receiving little information about their illness, its management, or appropriate self-care behaviors^6^. These challenges necessitate novel solutions aimed at mitigating the burden of HF, improving the patient experience, and offloading overburdened clinics.

Among the potential solutions to these challenges is the promotion and improvement of HF self-care. Such patient-centred care strategies aim to empower patients with skills and behaviours that will improve illness management and health outcomes^7^. Self-care interventions have been found to enable patients to better manage their own condition, and have been successfully implemented in other resource-limited settings^8–11^. Various technological and non-technological methods of improving chronic disease self-care have been investigated in low- and middle-income countries (LMICs), with digital health interventions appearing to be the most common modality^12^. Various factors make digital health interventions favourable for improving self-care in resource-limited settings, such as the near-ubiquity of mobile phone penetration in LMICs^13^, the ability to automate simple tasks with minimal oversight from oft-overburdened clinical staff, and the functionality to algorithmically triage patients in order to optimize resource allocation^14^. Despite the potential benefits of self-care tools, HF-specific self-care remains underutilized globally, with one of the greatest performance gaps coming from LMICs like Uganda^12^.

Our research team previously developed a digital health intervention (*Medly*) for use in Canada, which features a validated rules-based algorithm to triage patient symptoms, deliver tailored self-care advice to patients, and alert clinicians of signs of regression in patient health^15^. *Medly* has been shown to reduce HF-related hospitalizations and to improve patient self-care ability^16,17^. As an extension of these positive results, as well as the aforementioned need for improved HF self-care in LMICs, our team recently performed formative research with the goal of adapting *Medly* for use in the Ugandan context. This formative research included a scoping review of previous self-care interventions deployed amongst patients with non-communicable diseases in LMICs^12^, as well as a mixed-methods study to explore the lived experiences and technological literacy of HF patients and clinicians at a cardiac care centre in Uganda^18^. In the latter study, we concluded that a) Ugandan HF patients and clinicians could benefit from an intervention that empowers patients to better care for themselves, and b) that such an intervention should incorporate Unstructured Supplementary Service Data (USSD) technology that is already familiar to Ugandans as a result of its use in local mobile money services. Accordingly, we implemented a user-centred design (UCD) process^19^ to develop a USSD-based self-care intervention called *Medly Uganda*. The UCD process and the resulting digital health intervention are described in this manuscript.

## Methods

### Study Design

This study used a UCD process for the conceptualization, design and development of *Medly Uganda*. Design activities were based primarily at the Uganda Heart Institute (UHI), a cardiac care centre located in Kampala, Uganda. As a design philosophy, the UCD process directs the creation of technologies with consistent consideration of end-user needs throughout three main stages of design: ideation, design and development, and evaluation (**Figure 1**)^19^. Guided by this philosophy, our multidisciplinary team of patients, clinicians, researchers, engineers and designers worked to develop an intervention that was both effective and contextualized to the local population. All research activities were approved by the Uganda National Council of Science and Technology (HS 2364), the Makerere University School of Medicine Research and Ethics Committee (REF 2017–076), the University Health Network Research Ethics Board (ID# 14-7510), and the Yale Human Research Protection Program (ID# 2000025338).

**Figure 1:**
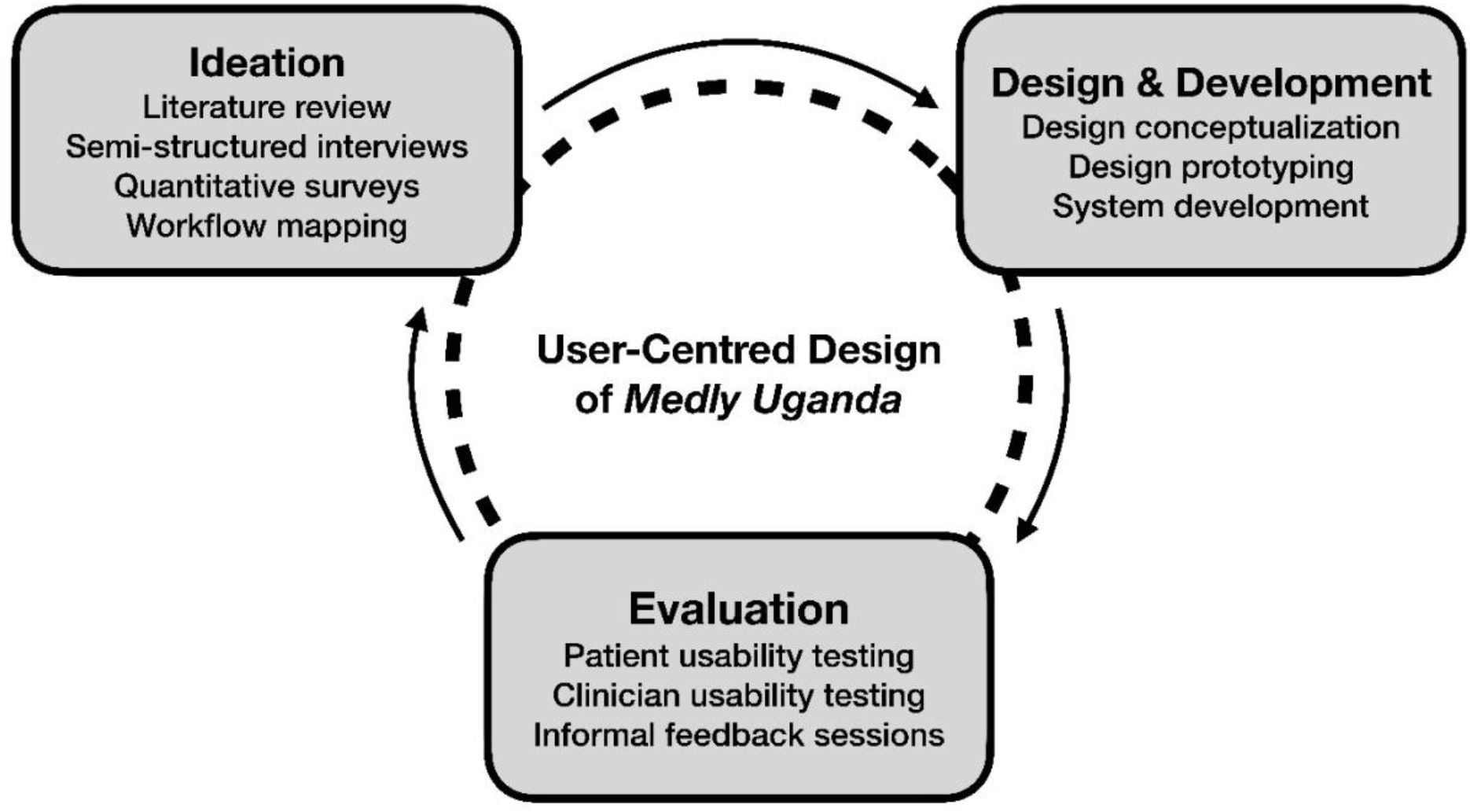
User-centred design framework (adapted from McCurdie et al.^19^)

### Phase 1: Ideation

The aims of the first phase of the UCD process were 1) to obtain a baseline understanding of the use of chronic disease self-care in LMICs, 2) to work with Ugandan HF patients and clinicians to define the specific needs and functionalities of a potential digital health intervention, and 3) to gain an understanding of the technical resources and on-the-ground personnel available to support the delivery and implementation of the intervention. We first conducted a systematic scoping review on the use of self-care for non-communicable diseases in LMICs, with the goal of summarizing the nature and effectiveness of past interventions that have enabled chronic disease self-care in resource-limited settings^12^. To better understand the HF-related needs and available technological resources in Uganda, we then conducted a mixed-methods study focused on the experiences and technological literacy of HF patients and clinicians at UHI^18^. Based on the key findings in this preliminary research, a list of design principles was defined to guide the development of a digital health intervention.

### Phase 2: Design and Development

The aim of the second phase of the UCD process was to utilize the elicited feedback from patients and clinicians to help design an intervention prototype. A preliminary design, consisting primarily of schematics and sketches describing the main components of the intervention, was created to allow for members of the multidisciplinary research team to provide feedback and suggest refinements to the system design over a series of iterations. Throughout this phase, we maintained consistent collaboration with Ugandan colleagues to allow for the system prototype to be effectively contextualized to the local setting. After several iterations during which refinements were made to the design prototype, we proceeded to develop the prototype into a fully-functional digital health intervention. During this design and development phase, our research team (based at the Uganda Initiative for Integrated Management of Non-Communicable Diseases) also developed strong relationships with key stakeholders in Uganda. Namely, the team fostered connections with UNICEF, the Uganda Ministry of Health, and a Ugandan technological consulting firm entitled GoodCitizen.

### Phase 3: Evaluation

The aim of the third phase of the UCD process was to evaluate the design of the digital health intervention to help ensure that the system could be effectively implemented in practice. Both HF patients and clinicians were invited to participate in usability testing of the patient- and clinician-facing components of the system, respectively. Participants were recruited from UHI. Patient eligibility criteria included a diagnosis of HF by a clinician at UHI and the ability to communicate in English, Luganda or Runyankole. Clinicians were considered eligible if they were a physician or nurse with direct experience caring for HF patients at UHI. At the start of each usability test, a high-level overview of the *Medly Uganda* system and the long-term vision for its deployment at UHI was provided. Participants were then asked to follow a series of simulated scenarios on their own device to test the functionality of the system. The goal of these scenarios was to capture the manner in which the participant interacted with the system in various situations, allowing for the isolation of key areas for improvement. Each session took place at UHI and lasted between 30 and 60 minutes for patient participants, and between 20 and 45 minutes for clinician participants. All participant interactions with the system were video recorded for subsequent analysis by the research team. For the patient interviews, an interpreter capable of speaking English, Luganda and Runyankole was present to allow patients to participate and interact with the system in their language of preference. Patients that were accompanied by a caregiver were given the option of a) using the system themselves, or b) having their caregiver use the system on their behalf.

## Results

### Phase 1: Ideation

Detailed results of both the scoping review and mixed-methods research amongst HF patients and clinicians at UHI are summarized elsewhere^12,18^. **Table 1** correlates the key findings that emerged in this formative research with the associated design principles that were formulated to guide the development of *Medly Uganda*.

**Table 1:** Summary of design principles for *Medly Uganda*.

| Research Finding | Design Principle |
| --- | --- |
| **Financial challenges:** Patients struggle with consultation, treatment, medication and travel costs. | The system must be free for the patients to use, and will ideally play a role in reducing existing costs associated with the HF condition. |
| **Fragmented self-care:** Patients demonstrate a lack of continuity in their self-care behaviours. Behaviours are instead often cyclical in nature. | The system will encourage continuous self-care behaviours, and educate the patients on the ill effects of lapses in self-care. |
| **Health dichotomy:** Patients use very extreme language when describing their state of health; often feeling as if they may “die tomorrow”. | The system must reassure patients that the HF condition can be managed. Patients must also be reminded that self-care behaviours are still necessary when they are feeling well. |
| **Limited access to medical devices:** Very few patients have regular access to a blood pressure monitor or weight scale. | The system must not depend on the use of medical devices. |
| **Need for information:** Patients have a limited understanding of their illness and self-care behaviours. Patients are also highly dependent on clinicians for health-related information. | The system must act as an alternative channel for disease-specific knowledge transfer and training of self-care behaviours, in turn reducing the demand on clinicians. |
| **Overburdened clinics:** Clinicians report a heavy workload, and note that this burden results in shorter interactions with patients. | The system will play a role in mitigating clinic burden by improving patient education and by reducing the number of unnecessary and/or emergency patient visits to the clinic. |
| **Regain of control:** Patients feel as if they lose control when experiencing symptoms, but express great relief upon receiving effective treatment. | The system will empower patients to take control of their health, and to maintain this sense of control in the midst of symptoms. |
| **Reverence for clinician:** Patients hold the clinician in extremely-high regard, and often report a sense of comfort and/or peace of mind after meeting with the clinician. | The system will improve communication between the clinician and patient, so that this sense of comfort can be achieved outside of a clinic visit. |
| **Technological literacy:** Patients report a high level of comfort with calling and Unstructured Supplementary Service Data (USSD)-based platforms, and less confidence with sending short message service messages and interacting with interactive voice response systems. | The system will harness the calling and USSD functionalities of the patient’s mobile device to maximize usability. |

### Phase 2: Design and Development

#### Overview of Medly Uganda

Guided by the design principles and consistent feedback from a multidisciplinary team, an iterative process was used to develop, refine and contextualize a digital health intervention named *Medly Uganda*. The digital health intervention is accessible by HF patients using any mobile device (smartphone or non-smartphone). Patients can dial into *Medly Uganda* using a USSD code and subsequently respond to a series of yes-or-no questions pertaining to their HF symptoms. Patients are asked to complete this symptom questionnaire on three pre-specified mornings per week. Upon receiving the patient’s symptom data, the system auto-generates self-care advice that is immediately sent to the patient via a short message service (SMS) message. For patients reporting symptoms in keeping with potential decompensation, the system also sends an SMS message to a reporting clinician containing a brief summary of the patient’s symptoms and a number at which the patient can be reached. The clinician can then review the patient’s complete history in a web-based dashboard, either on their computer or smartphone. After reviewing the patient’s file, the clinician can call the patient to discuss their symptoms and subsequently decide whether a visit to the clinic is necessary, or whether the patient’s issue can be managed remotely. **Figure 2** offers a high-level overview of the *Medly Uganda* system.

**Figure 2:**
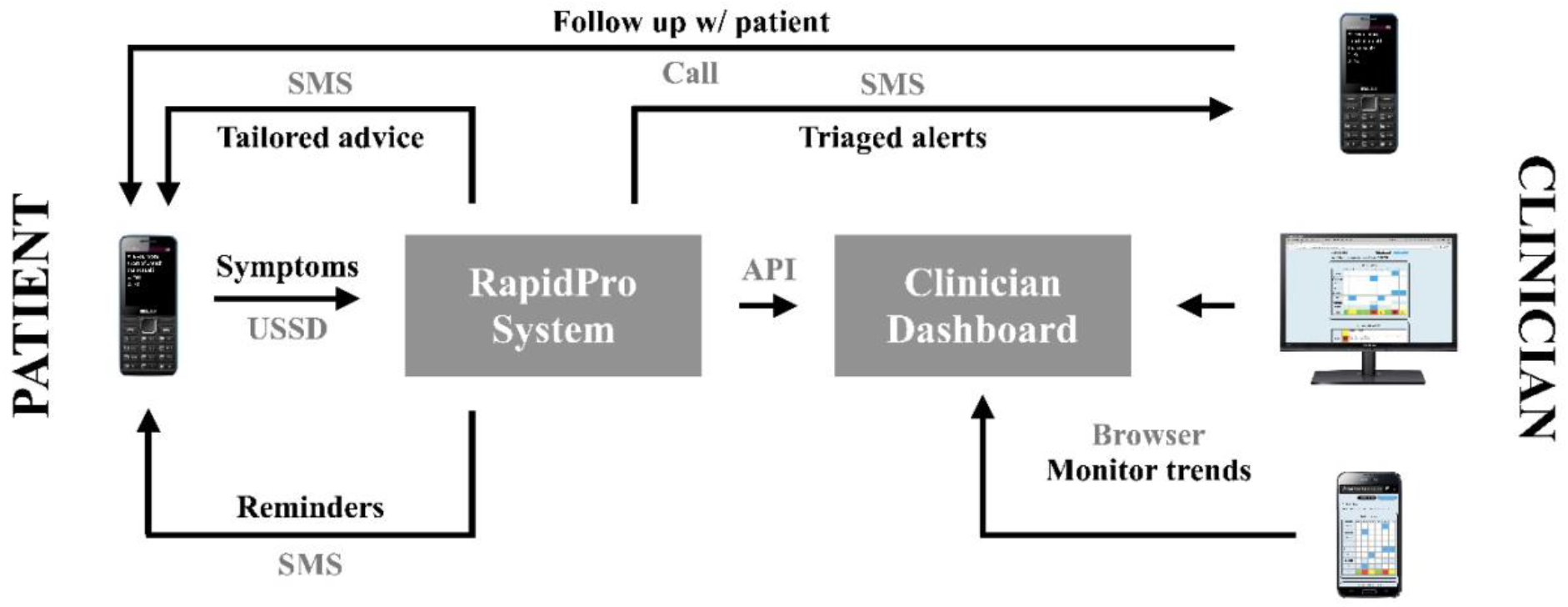
Overview of *Medly Uganda*.

#### Patient-Facing System

The patient-facing portion of *Medly Uganda* involves a symptom questionnaire and tailored self-care advice, both of which are guided by an inbuilt algorithm. The decision-tree algorithm was designed to take responses to a symptom questionnaire as input, and return appropriate self-care advice based on the inputted symptoms. The algorithm also uses the reported symptoms to triage the patient with a current disease state. The algorithm was initially based on the *Medly* system currently deployed in Canada^17^, with modifications being made to contextualize the algorithm based on feedback from our Ugandan colleagues. After applying an iterative design process, the final algorithm includes seven yes-or-no symptom questions and six potential disease states. Based on symptom responses, patients can be triaged as either *low risk, moderate risk, moderate risk with fluid overload, high risk, high risk with fluid overload*, or *critical*. Each of these disease states corresponds to a specific self-care message that is sent to the patient. Moreover, moderate- and high-risk disease states trigger a message that is sent to the clinician for potential follow-up.

To access the symptom questionnaire, patients dial a USSD code on their mobile device (e.g. *123#). Upon accessing the system, patients are asked to enter a four-digit password that they set during their registration to the system. Upon entering the correct password, the user is presented with a series of symptom-related questions in the language that they selected during the registration process (i.e. English, Luganda or Runyankole). In situations where a patient forgets their password, they are able to contact technical support to reset their password. Similar to USSD-based mobile money services commonly used in Uganda, patients can then respond to the yes-or-no questions using the keypad on their device. Specifically, patients are asked to respond with “1” for *Yes*, “3” for *No*, or “9” to repeat the previous question. The selection of numeric responses that are physically separated on the keypad (e.g. 1, 3 and 9) minimizes the risk of an accidental entry of the wrong response. Once the patients complete the questionnaire, the USSD session is terminated and an SMS message is sent to their device containing tailored self-care advice. **Figure 3** demonstrates two screens in the USSD workflow, as well as the resulting SMS message.

**Figure 3:**
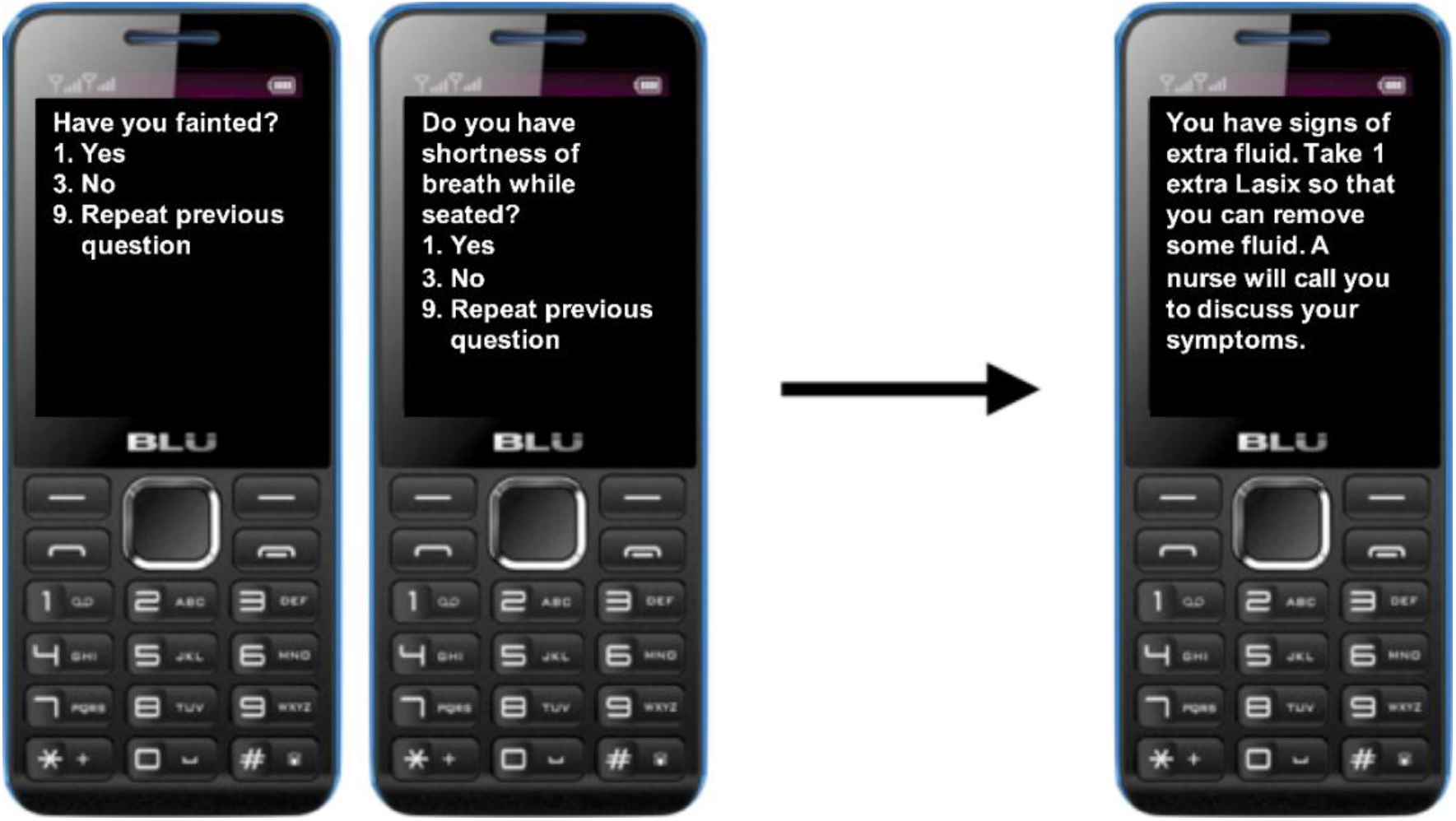
Simulated screenshots of the USSD workflow and resulting SMS message.

To operationalize the above interaction between the patient and *Medly Uganda*, an open-sourced software entitled RapidPro was used. RapidPro was chosen over similar platforms due to the ease with which modifications could be made to the system, its compatibility with Ugandan USSD channels, and its previous use by collaborators at UNICEF and the Uganda Ministry of Health for their USSD-based *FamilyConnect* system^20^ into which *Medly Uganda* was being integrated. Using RapidPro, a digital health system was developed comprising two components. An interaction with the system begins with a USSD workflow (i.e. the *symptom flow*), where patient responses to the symptom questionnaire and the developed decision-tree algorithm are used to determine the patient’s disease state on a given day. Completion of the USSD flow then triggers the start of an SMS workflow (i.e. the *message flow*), where the system uses the determined patient disease state to send the appropriate SMS message(s) to the patient and clinician (if required). A high-level overview of this workflow is pictured below in **Figure 4**.

**Figure 4:**
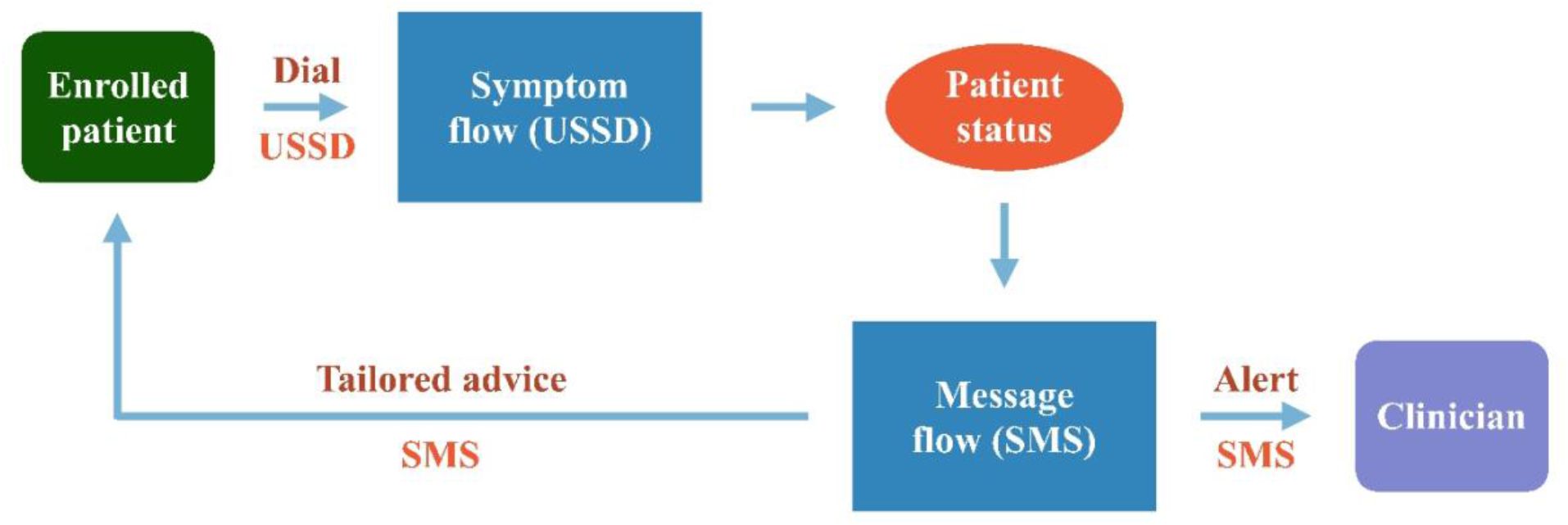
Overview of RapidPro system.

A final consideration that is integrated into the patient-facing system is adherence reminders. Instead of prompting every patient three times per week, the RapidPro system automatically sends SMS reminders to patients who have not reported their symptoms by midday on days on which a symptom report is expected.

#### Clinician-Facing System

The clinician-facing system comprises autogenerated SMS alerts sent directly to the clinician’s mobile device, as well as a web-based dashboard accessible from the clinician’s smartphone or computer. The SMS alerts are triggered in response to a patient-submitted symptom report that is triaged as either moderate or high risk. These SMS messages include a brief overview of the patient’s symptoms, as well as a phone number at which the patient can be reached. The phone number can also be used to query the patient’s file in the web-based dashboard.

The web-based dashboard is a simple and secure platform that enables clinicians to access additional information relating to their presenting patient. The web application uses React JavaScript library for the front end, Express JavaScript library for the back end, and PostgreSQL for the database. The application is hosted using the cloud platform Heroku, while the PostgreSQL database is stored on a cloud server hosted by UHI. Clinicians access the web application by navigating to a UHI-hosted web address on their internet-enabled device. After entering their credentials on the *Login* page (**Figure 5A**), the clinician is provided with the option of querying their patient using a phone number, browsing all of their patients by name, or adding a patient to the system (**Figure 5B**). Adding a new patient requires simple demographic information including a name, phone number, date of birth, sex, and preferred language (**Figure 5C**). A clinician may also choose to add credentials that will allow another clinician to access the system (**Figure 5D**). If the clinician queries a patient using the patient’s phone number, they will be taken directly to the patient’s file. If they choose to browse all of their patients, they are taken to the *Search Patient by Name* page where they can search patients using their first name, last name or phone number (**Figure 5E**).

**Figure 5:**
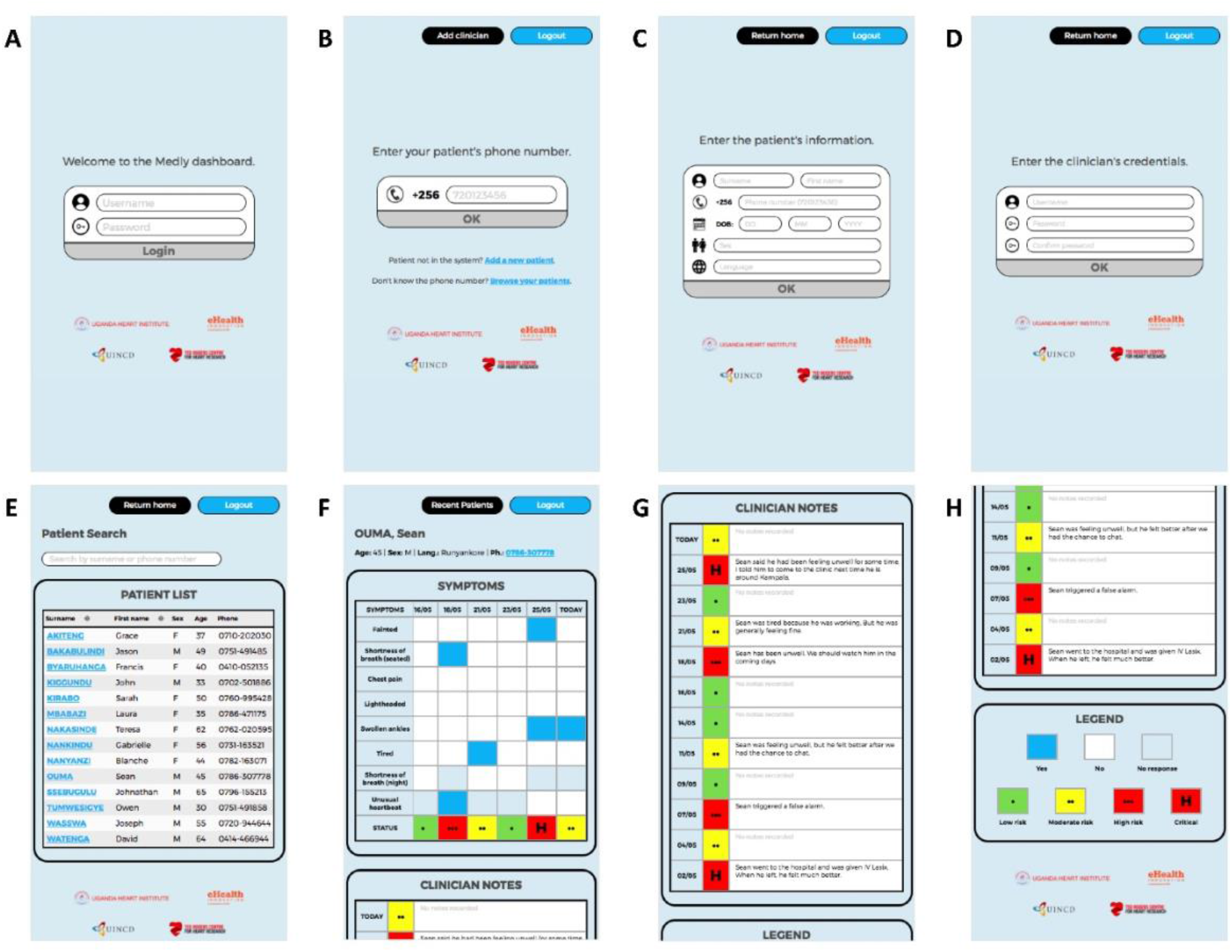
User interface for clinician dashboard as seen on a smartphone.

Once the clinician has accessed a specific patient file, they will see simple demographic information at the top of the page, a panel summarizing the patient’s symptom history (**Figure 5F**), a panel including clinician notes relating to past symptom reports (**Figure 5G**), and a legend for the symbols used in the symptom history panel (**Figure 5H**). The symptom history is presented as a block chart, with the y-axis representing the various items included in the symptom questionnaire (e.g. chest pain, fainting) and the x-axis indicating the report dates. For a specific symptom question on a given date in the chart, a blue square denotes a “Yes” response, a light gray square signifies a “No” response, and a dark gray square indicates that the corresponding symptom question was not asked on the specific date. In addition to the symptom responses, the patient’s assigned status for each report is included at the bottom of the chart. Risk status is represented using a *stoplight* model already used by clinicians at UHI (i.e. green for low risk, yellow for moderate risk, red for high risk), with an “H” being added to critical reports where the patient has been instructed to report to a hospital. To increase accessibility of the dashboard to clinicians with colorblindness, risk status is also denoted with an increasing number of dots (i.e. 1 for low risk, 2 for moderate risk, 3 for high risk). Below the symptom history, clinicians are provided with a space to add notes for each symptom report submitted by the patient. These note fields save automatically as the clinician types, further increasing the usability of the dashboard.

### Phase 3: Evaluation

#### Patient Usability Testing

In-person usability tests were performed with six HF patients and four informal caregivers at UHI. Six participants opted to use the system in Luganda, whereas four participants completed the testing in English. No users performed usability testing in Runyankole. Overall, participants were extremely satisfied with both the usability and usefulness of the intervention. There was strong agreement amongst patients that the system was straightforward, easy to learn and contextual to the user. Participants appreciated the similarities between the developed system and mobile money services with which they were already familiar. The patients also exhibited a strong interest in future use of the system as part of their daily self-care regimen. Even patients who self-identified as having low technological literacy were able to learn and use the system. For these participants, the research team allowed them to experiment with submitting different sets of symptoms until they reached a point where they felt confident that they could use the system independently at home. By the end of the usability testing, every participant indicated that they were able to reach this level of comfort with the system.

The usability testing allowed for the identification of key areas for further improvement. Most notably, the usability tests highlighted the importance of appropriate training when patients are first introduced to the system. Certain individuals, particularly those that opted to use the system in Luganda, initially had difficulty completing the USSD interaction in the allotted time. These patients often took increased time to read, comprehend and respond to the questions posed by the system. After a brief introductory discussion with the research team regarding each question, however, these patients quickly adjusted to the time requirements of the system. Moreover, several patients reported that they would not know which pill to take when advised by the system to take an extra diuretic (i.e. “Lasix”) pill. Based on this confusion, the initial training was further modified to include a brief discussion of the patient’s HF medications and how specifically to identify the diuretic pill.

#### Clinician Usability Testing

Usability tests were also performed with five cardiologists and two nurses. Clinicians reported a high degree of usability, simplicity and consistency for the dashboard. Participants also described the system as straightforward and easy to learn. Following a brief overview of the dashboard by the research team, clinicians quickly mastered the dashboard and its various functions. Based on the time taken to complete the typical interaction of logging in, querying a patient, reviewing the patient’s history, and subsequently recording a note, it was estimated that responding to a single patient alert could take as little as two-to-four minutes of in-dashboard interaction (i.e. not including clinician-patient telephone communication).

Qualitative assessment of the clinician usability tests helped identify important usage patterns and elements of the dashboard that limited usability. For example, clinicians would often affix a leading zero when entering a phone number into the dashboard (e.g. 0720123456); thus, the dashboard was updated to omit zeros located at the beginning of a phone number. Other minor issues included the requirement to click a patient’s last name to access their file from the *Browse Patients by Name* screen, rather than the expected functionality of clicking the row associated with their name. Again, the dashboard was modified to accommodate the expected functionality.

## Discussion

This paper presents the user-centred design and development of a digital health intervention, *Medly Uganda*, for HF patients and clinicians living in Uganda. *Medly Uganda* is designed to empower HF patients to improve their HF self-care through a simple symptom-focused questionnaire, an autonomous triage algorithm, and self-care advice sent directly to the patient’s device. Moreover, the system supports the remote management of HF patients by alerting clinicians to moderate- or high-risk patients and enabling clinicians to review patient information in a web-based dashboard.

The strengths of the presented intervention are numerous. The user-centred design process enabled the development of a contextualized system designed specifically to address patient- and clinician-raised concerns regarding HF management in Uganda. This end-user input informed several important design characteristics, such as the use of the USSD platform, the focus on addressing information needs amongst patients, and the semi-autonomous nature of the eventual design. The involvement of Ugandan HF patients and clinicians at all stages of design and development will also help support the adoption, acceptability and scalability of *Medly Uganda* in the long term.

Several potential benefits are also associated with the autonomous triage algorithm. The autonomous nature of the algorithm will enable clinics to efficiently manage a larger number of HF patients. By autonomously triaging patients in terms of their risk status, clinics can prioritize the care of high-risk patients while continuing to remotely monitor individuals that remain asymptomatic. This triage system presents potential benefits towards clinic operation, as it should reduce the number of unnecessary visits to the clinic and increase availability for patients with acute concerns. An additional strength of the algorithm is the potential impact that the resulting self-care advice will have on patient quality of life. When patients are triaged as low risk, they will receive positive reinforcement to maintain their current self-care behaviours. When a patient has signs of decompensation, they will be reassured through tailored self-care advice and timely intervention by their care provider (if required).

An additional strength of *Medly Uganda* is that the patient-facing portion of the system harnesses a technology that is both familiar and accessible to Ugandan HF patients. The USSD platform is ubiquitous in Uganda as a result of its use in local mobile money services. Our formative research amongst the HF population at UHI found that a significantly larger proportion of patients identified as comfortable using their phone for mobile money services when compared to both SMS-based services and interactive voice response systems^18^. Because USSD is accessible on any network-enabled device (smartphone or non-smartphone), the platform is accessible to individuals with varied economic means. USSD also enables the system to be provided to patients free of charge, as USSD-associated costs are incurred by the service provider. Accordingly, the USSD-based design increases both the familiarity and accessibility of the intervention to HF patients in Uganda.

A final strength of the design is the sustainable and collaborative approach that was used in all phases of development. In addition to the aforementioned involvement of HF patients and clinicians, early stakeholder engagement with UNICEF and the Uganda Ministry of Health provided crucial support in the development and deployment of the intervention. As UNICEF had already deployed a government-endorsed USSD-based system for pregnant and postpartum women entitled *FamilyConnect* (based on the South African *MomConnect*)^20^, we were able to leverage a breadth of experience and infrastructure to realize *Medly Uganda*. Moreover, the ongoing collaboration with UHI, Makerere University and the Uganda Ministry of Health will help to further solidify the long-term acceptability of *Medly Uganda* amongst key stakeholders.

Despite its promise, the *Medly Uganda* system is not without limitations. In its current form, the developed system requires patients to have access to a mobile device. Though our preliminary research indicated that 98% of patients at UHI have access to a mobile device^18^, we acknowledge that the small minority of patients without a mobile phone will be unable to access the system. Access may also be limited by patient literacy levels and technological capabilities. Our formative research suggested that nearly 30% of patients at UHI self-identify as illiterate;^18^ however, we attempted to mitigate the effect of this limitation by offering the service in three local languages. Similarly, we attempted to lessen the impact of technological illiteracy by opting for the USSD-based system preferred amongst patients at UHI^18^. An additional limitation is the time restrictions placed on a USSD interaction, which can vary between 90 and 180 seconds depending on the cellular provider. Though we expect that most patients will be able to complete the seven-item questionnaire in the allotted time, this time constraint could prohibit some patients from effectively submitting their symptoms. A final potential limitation of the system is that responding to patient alerts could eventually become burdensome for the assigned clinician(s) as the number of patients using *Medly Uganda* increases. We expect that the semi-autonomous design should mitigate this final limitation; however, it is possible that designated staff members will eventually be required to remotely manage the *Medly Uganda* users. Such implementation and service design considerations will be addressed in upcoming phases of validation and deployment.

*Medly Uganda* is currently being tested in a pilot hybrid implementation/clinical effectiveness trial among HF patients and clinicians at UHI. This pilot study will shed light on technical and human resource challenges involved in deploying and managing the system. It is important to recognize that we have designed the *Medly Uganda* system alongside a population of patients based at UHI, where access to care is relatively more available than more-remote regions across Uganda. Accordingly, our future work will build upon the results of this pilot trial with the goal of broadening the geographic and sociodemographic reach of the system through further collaborations with nursing stations throughout Uganda. As a long-term goal, we also hope to expand the scope of chronic conditions that can be managed using *Medly Uganda*.

To conclude, Ugandan HF patients and clinicians stand to benefit from digital health interventions that enable patients to better care for themselves. We used a UCD process to develop a USSD-based self-care intervention named *Medly Uganda* that allows HF patients to report daily symptoms, receive tailored treatment advice, and connect with a clinician when showing signs of decompensation. Usability testing showed *Medly Uganda* to be well-accepted by clinicians, patients and caregivers. As we move towards deployment of *Medly Uganda*, we expect the system to help decongest cardiac clinics and improve self-care efficacy amongst HF patients in Uganda.

## Data Availability

All authors had access to all data and had a role in writing the manuscript.

## Acknowledgements

We would like to acknowledge the patients, caregivers and clinicians that participated in the codevelopment of *Medly Uganda*. We would also like to thank Rebecca Gillman for her thoughtful review of this manuscript.

## Author Contributions

**JH** led the design, development and evaluation of *Medly Uganda*, as well as co-authored the original manuscript. **SW** co-authored the original manuscript. **PB** helped conduct usability testing and offered interpretation services. **JAC, IS, ARA, HJR** and **JIS** supervised all aspects of the study from ideation to evaluation. **JAC, HJR** and **JIS** acquired funding necessary to complete this study. **ES** provided insight throughout the design and development phases. **All authors** reviewed the manuscript and provided critical feedback.

